# Biomarkers of endothelial dysfunction and outcomes in coronavirus disease 2019 (COVID-19) patients: a systematic review and meta-analysis

**DOI:** 10.1101/2021.01.24.21250389

**Authors:** Andrianto, Makhyan Jibril Al-Farabi, Ricardo Adrian Nugraha, Bagas Adhimurda Marsudi, Yusuf Azmi

## Abstract

**Background:** Several studies have reported that the severe acute respiratory syndrome coronavirus 2 (SARS-CoV-2) can directly infect endothelial cells, and endothelial dysfunction is often found in severe cases of coronavirus disease 2019 (COVID-19). To better understand the pathological mechanisms underlying endothelial dysfunction in COVID-19-associated coagulopathy, we conducted a systematic review and meta-analysis to assess biomarkers of endothelial cells in patients with COVID-19.

**Methods:** A literature search was conducted on online databases for observational studies evaluating biomarkers of endothelial dysfunction and composite poor outcomes in COVID-19 patients.

**Results:** A total of 1187 patients from 17 studies were included in this analysis. The estimated pooled means for von Willebrand Factor (VWF) antigen levels in COVID-19 patients was higher compared to healthy control (306.42 [95% confidence interval (CI) 291.37-321.48], p<0.001; I^2^:86%), with the highest VWF antigen levels was found in deceased COVID-19 patients (448.57 [95% CI 407.20-489.93], p<0.001; I^2^:0%). Meta-analysis showed that higher plasma levels of VWF antigen, tissue-type plasminogen activator (t-PA), plasminogen activator inhibitor-1 antigen (PAI-1) antigen, and soluble thrombomodulin (sTM) were associated with composite poor outcome in COVID-19 patients ([standardized mean difference (SMD) 0.74 [0.33-1.16], p<0.001; I^2^:80.4%], [SMD 0.55 [0.19-0.92], p=0.003; I^2^:6.4%], [SMD 0.33 [0.04-0.62], p=0.025; I^2^:7.9%], and [SMD 0.55 [0.10-0.99], p=0.015; I^2^:23.6%], respectively).

**Conclusion:** The estimated pooled means shows increased levels of VWF antigen in COVID-19 patients. Several biomarkers of endothelial dysfunction, including VFW antigen, t-PA, PAI-1, and sTM, are significantly associated with increased composite poor outcome in patients with COVID-19.

**PROSPERO registration number:** CRD42021228821

## Introduction

Although initially recognized as a disease affecting the respiratory system, the coronavirus disease 2019 (COVID-19) often manifests as cardiovascular complications such as myocarditis, myocardial injuries, arrhythmias, and venous thromboembolism events (VTE) [1]. There are several possible mechanisms for these phenomena, one of which may be an unrestrained and unbalanced innate immune response, which in turn negates effective adaptive immunity, supporting the progression of COVID-19. Frequent laboratory abnormalities in patients with unfavorable progression of COVID-19 include abnormal cytokine profiles, which led to the initial presumption that the immune response to severe acute respiratory syndrome coronavirus 2 (SARS-CoV-2) infection involved a cytokine storm [2]. However, recent evidence suggests that increased inflammatory cytokines including interleukin-6 (IL-6) in patients with severe and critical COVID-19 are significantly lower compared to patients with sepsis and acute respiratory distress syndrome (ARDS) not associated with COVID-19, thus doubting the role of a cytokine storm in COVID-19-related multiple organ damage [3].

Several studies have reported that the SARS-CoV-2 can directly infect endothelial cells, and endothelial dysfunction is often found in severe cases of COVID-19 [4]. Autopsy findings have also demonstrated endothelial dysfunction and microvascular thrombosis, together with pulmonary embolism (PE) and deep-vein thrombosis in COVID-19 patients [5]. These findings suggest that endothelial injury, endotheliitis, and microcirculatory dysfunction in different vascular beds contribute significantly to life-threatening complications of COVID-19, such as VTE and multiple organ involvement [6]. To better understand the pathological mechanisms underlying endothelial dysfunction in COVID-19-associated coagulopathy, we conducted a systematic review and meta-analysis to assess biomarkers of endothelial cells in patients with COVID-19.

## Methods

### Search strategy and study selection

A systematic literature search of PubMed, PMC Europe, and the Cochrane Library Database from 1 January 2020 to 20 December 2020 was performed using the search strategy outlined in Supplementary Table S1. Additional records were retrieved from Google Scholar. Duplicate articles were removed after the initial search. Two authors (MJA and YA) independently screened the title and abstract of the articles. Articles that passed the screening were assessed in full text based on the eligibility criteria. Disagreements were resolved by discussion with the senior author (A). This study was conducted following the Preferred Reporting Item for Systematic Reviews and Meta-Analysis (PRISMA) statement and registered with the International Prospective Register of Systematic Reviews (PROSPERO) database (registration number CRD42021228821).

### Eligibility criteria

We included all observational studies examining biomarkers of endothelial dysfunction and outcomes from patients who tested positive for SARS-CoV-2 using the reverse transcription-polymerase chain reaction (RT-PCR) test. The following types of articles were excluded from the analysis: case reports, review articles, non-English language articles, research articles on the pediatric population, animal or in-vitro studies, unpublished studies, and studies with irrelevant or non-extractable results.

### Data collection process

Data extraction was carried out by two authors (MJA and YA) independently using piloted data extraction forms consisting of the author, year of publication, study design, number and characteristics of samples, levels of several biomarkers of endothelial dysfunction, and patient outcomes. The biomarkers of endothelial dysfunction analyzed in this study were von Willebrand Factor (VWF) antigen, tissue-type plasminogen activator (t-PA), plasminogen activator inhibitor-1 antigen (PAI-1), and soluble thrombomodulin (sTM). The primary endpoint was composite poor outcomes consisting of ICU admission, severe illness, worsening of the respiratory status, the need for mechanical ventilation, and mortality. Moreover, if the included studies reported the data using median and quartile values, we used the formula developed by Wan *et al*. to estimate mean and standard deviation [7]. Disease severity was defined based on the WHO R&D Blueprint on COVID-19 [8].

### Quality assessment

The quality and risk of bias assessment of included studies were conducted using the Newcastle-Ottawa score (NOS) [9] by all authors independently, and discrepancies were resolved through discussion. This scoring system consists of three domains: the selection of sample cohorts, comparability of cohorts, and assessment of outcomes (Supplementary Table S2).

### Data analysis

Stata software V.14.0 (College Station) was used for meta-analysis, and figure of estimated pooled means were produced using GraphPad Prism 9. We pooled multiple means and standard errors of the same population characteristic from different studies into a single group using the fixed-effect model of meta-analysis algorithm. Pooled effect estimates of the outcomes were reported as standardized means differences (SMD). Fixed-effects and random-effects models were used for pooled analysis with low heterogeneity (I^2^ statistic <50% or P-value <0.1) and high heterogeneity (I^2^ statistic> 50% or P-value ≤ 0.1), respectively. Statistical significance was determined with P-value < 0.05. We performed a sensitivity analysis to test the robustness of the pooled effect estimates for VWF antigen levels by excluding each study sequentially and rerunning the meta-analysis. The funnel-plot analysis was used to assess the symmetrical distribution of effect sizes, and the regression-based Egger test was performed to assess publication bias on continuous endpoints.

## Results

### Study characteristics

We identified 697 articles from the database search, and 492 articles remained after the duplication was removed. Screening on titles and abstracts excluded 465 articles, and the remaining 27 full-text articles were assessed according to eligibility criteria. As a result, 17 studies [10–26] with a total of 1187 patients were subjected to qualitative analysis and meta-analysis (Figure 1 and Table 1). Quality assessment with NOS showed that 13 studies [10,13– 19,21–24,26] were of good quality with ≥ 7 NOS score and four studies [11,12,20,25] were considered as moderate quality with six NOS score (Supplementary Table S2).

**Table 1.** Characteristics of the included studies.

| No | Study | Population | Number of samples | Age mean (SD) | Male number (%) | Comparison / end point measure | Marker examined | NOS |
| --- | --- | --- | --- | --- | --- | --- | --- | --- |
| 1 | Cugno, 2020 [10] | All hospitalized patients | 148 | 61 (13) | 87 (59) | ICU admission | VWF antigen, sTM, PAI-1 antigen, t-PA | 8 |
| 2 | Fan, 2020 [11] | ICU patients | 12 | 51 (17) | 11 (92) | NA | VWF antigen | 6 |
| 3 | Goshua, 2020 [19] | All hospitalized patients | 68 | 62 (16) | 41 (60) | ICU admission | VWF antigen, sTM, PAI-1 antigen | 9 |
| 4 | Helms, 2020 [20] | ICU patients | 150 | 62 (14) | 122 (81) | NA | VWF antigen | 6 |
| 5 | Henry, 2020 [21] | All hospitalized patients | 52 | 52 (21) | 30 (58) | Severe COVID-19 | VWF antigen | 9 |
| 6 | Hoechter, 2020 [22] | ICU patients | 22 | 62 (14) | 19 (86) | NA | VWF antigen | 7 |
| 7 | Ladikou, 2020 [23] | ICU patients | 24 | 64 (13) | 18 (75) | ICU admission | VWF antigen | 7 |
| 8 | Mancini, 2020 [24] | All hospitalized patients | 50 | 58 (13) | 32 (64) | ICU admission | VWF antigen | 7 |
| 9 | Morici, 2020 [25] | ICU patients | 6 | 62 (5) | 4 (67) | NA | VWF antigen | 6 |
| 10 | Nougier, 2020 [26] | All hospitalized patients | 78 | 60 (14) | 51 (65) | ICU admission | PAI-1 antigen, t-PA | 9 |
| 11 | Panigada, 2020 [12] | ICU patients | 24 | 56 (15) | NA | NA | VWF antigen | 6 |
| 12 | Pine, 2020 [13] | All hospitalized patients | 49 | 63 (17) | 33 (67) | ICU admission | PAI-1 antigen | 8 |
| 13 | Ranucci, 2020 [14] | ICU patients | 20 | 64 (12) | 16 (80) | Mortality | t-PA | 8 |
| 14 | Rauch, 2020 [15] | All hospitalized patients | 243 | 64 (16) | 155 (64) | Worsening of the respiratory status | VWF antigen | 7 |
| 15 | Rovas, 2020 [16] | All hospitalized patients | 23 | 64 (17) | 20 (87) | Mechanical ventilator | sTM | 9 |
| 16 | Sweeney, 2020 [17] | All hospitalized patients | 181 | 66 (15) | 106 (59) | Mortality | VWF antigen | 7 |
| 17 | Taus [18] | All hospitalized patients | 37 | 62 (13) | 18 (49) | NA | VWF antigen, VWF activity | 8 |

ICU, intensive care unit; NA, not available; VWF, von Willebrand Factor; PAI-1, plasminogen activator inhibitor-1 antigen; sTM, soluble thrombomodulin; t-PA, tissue-type plasminogen activator; NOS, Newcastle-Ottawa Scale.

**Figure 1.**
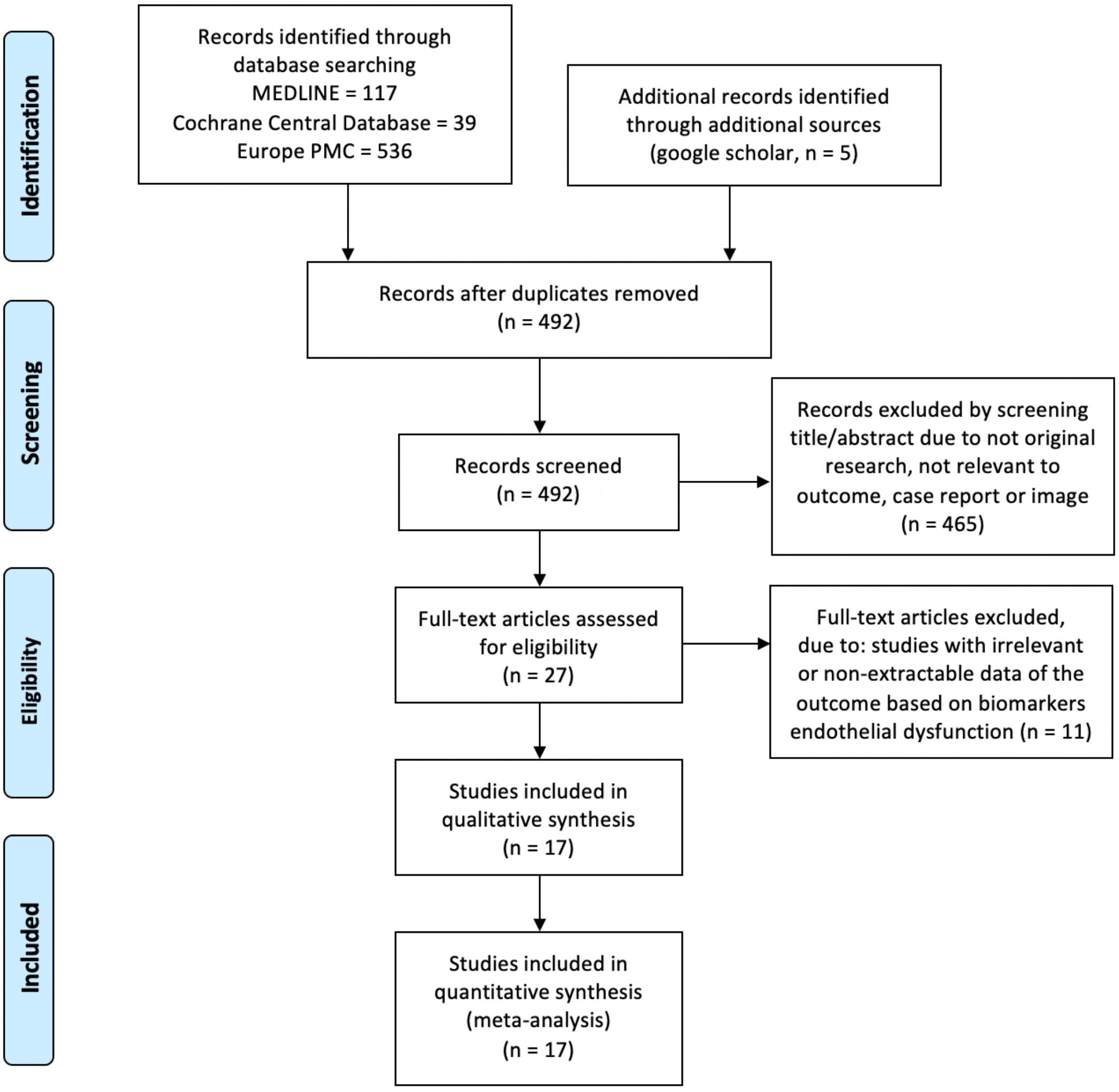
PRISMA flowchart.

### Biomarkers of endothelial dysfunction and outcome

There was an increase in the VWF antigen levels in COVID-19 patients with different levels for each outcome group of COVID-19 patients. The pooled means plasma levels of VWF antigen in COVID-19 patients treated at the general wards and ICU patients or severely ill patients ([306.42 [95% confidence interval (CI) 291.37-321.48], p<0.001; I^2^:86%] and [398.56 [95% CI 386.84-410.30], p<0.001; I^2^:92%], respectively) were higher than healthy controls (103.24 [95% CI 91.31-115.17], p<0.001; I^2^:0%). Moreover, deceased COVID-19 patients had the highest pooled means of VWF antigen levels (448.57 [95% CI 407.20-489.93], p<0.001; I^2^:0%) (Figure 2).

**Figure 2.**
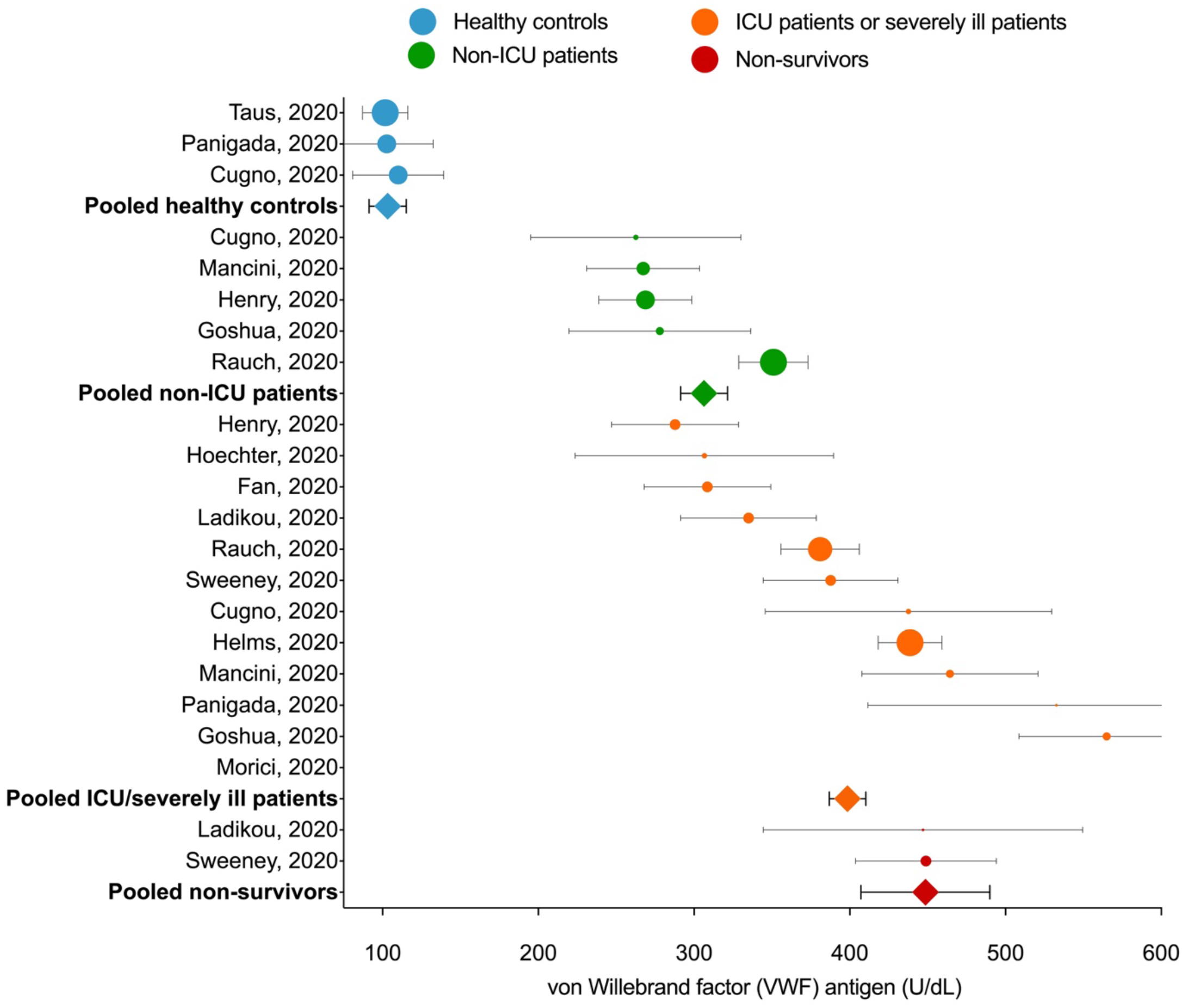
The estimated pooled mean for plasma levels of Von Willebrand Factor antigen in patients with COVID-19. For individual studies, circle markers indicate study means and error bars indicate 95% confidence intervals. Markers are sized proportionately to the weight of the study in the analysis. Estimated pooled means for grouped studies are represented by the square markers.

Meta-analysis showed that higher plasma levels of VWF antigen was associated with composite poor outcome (SMD 0.74 [0.33-1.16], p<0.001; I^2^:80.4%). Patients with poor outcome had significantly a higher level of t-PA and PAI-1 antigen compared to patients with good outcomes ([SMD 0.55 [0.19-0.92], p=0.003; I^2^:6.4%] and [SMD 0.33 [0.04-0.62], p=0.025; I^2^:7.9%], respectively). The plasma levels of sTM were found to be higher in COVID-19 patients with poor outcome ([SMD 0.55 [0.10-0.99], p=0.015; I^2^:23.6%]) (Figure 3).

**Figure 3.**
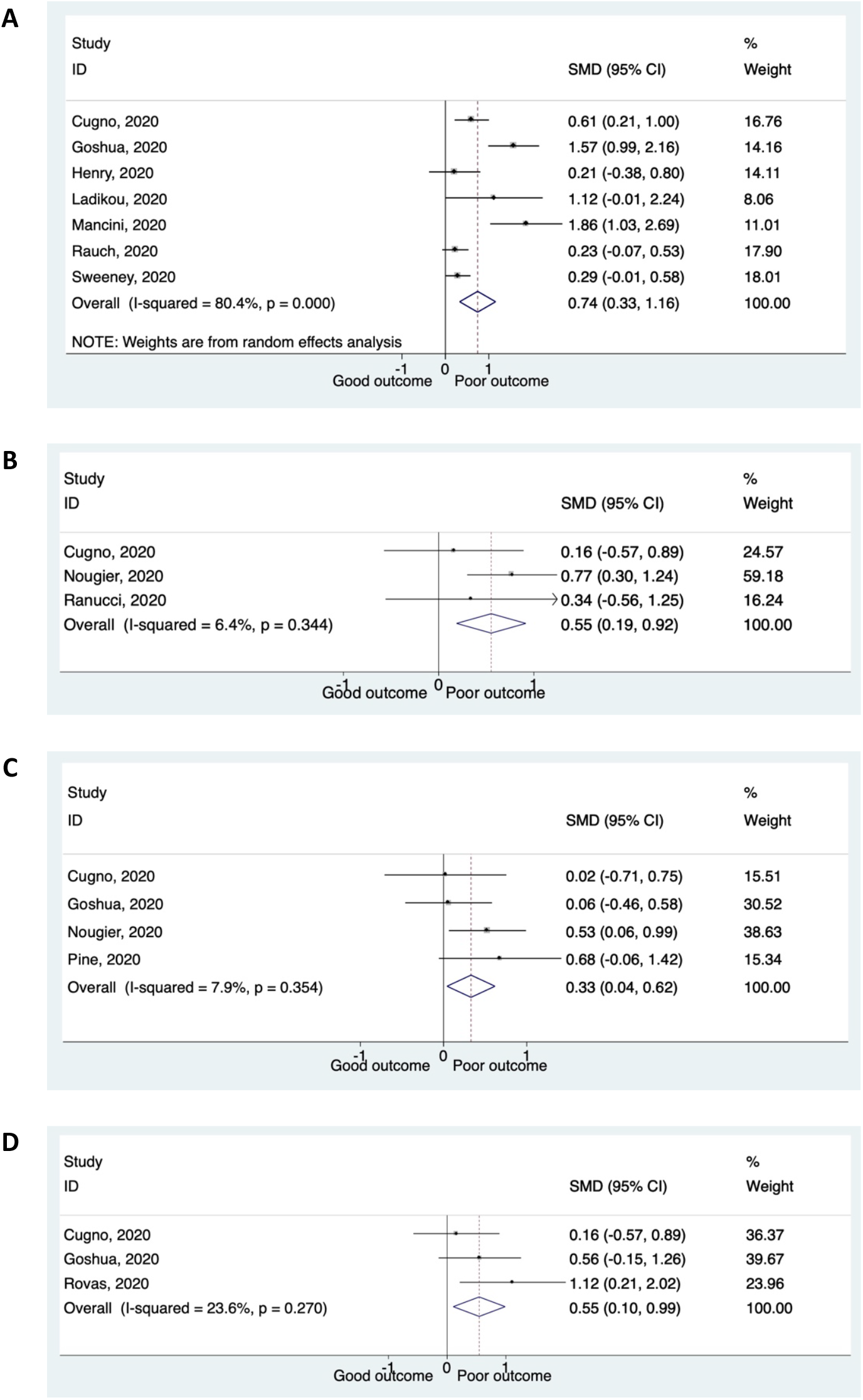
Several biomarkers for endothelial dysfunction and the outcome of COVID-19. Patients presenting with a higher plasma levels of (A) von Willebrand Factor (VWF) antigen; (B) tissue-type plasminogen activator (t-PA); (C) plasminogen activator inhibitor-1 antigen (PAI-1) antigen; and (D) soluble thrombomodulin (sTM) have an increased risk of composite poor outcome.

We found substantial heterogeneity for VWF antigen analysis (I^2^:80.4%) and low heterogeneity for t-PA, PAI-1 antigen, and sTM analysis (I^2^:6.4%, I^2^:7.9%, and I^2^:23.6%, respectively). However, subgroup analyses to evaluate potential sources of heterogeneity of VWF levels were not performed due to the small amount of primary data included in the group analysis. The sensitivity analysis of VWF levels after excluding two studies [19,24] at risk of bias decreased the heterogeneity considerably while maintaining the significance of pooled effect estimate (SMD 0.34 [0.17-0.62], p <0.001; I2:9.2%).

### Publication bias

The visual assessment of the funnel plot showed an asymmetrical shape for the analysis of the vWF antigen levels, which indicated the possibility of publication bias (Figure 4). This asymmetrical shape was due to the inclusion of the studies by Goshua *et al*. [19] and Mancini *et al*. [24]. However, quantitative analysis using regression-based Egger’s test for the same variable showed no significant result of small-study effects (p=0.063). Regression-based Harbord’s test for other biomarkers and composite poor outcome also showed no significant result of small-study effects.

**Figure 4.**
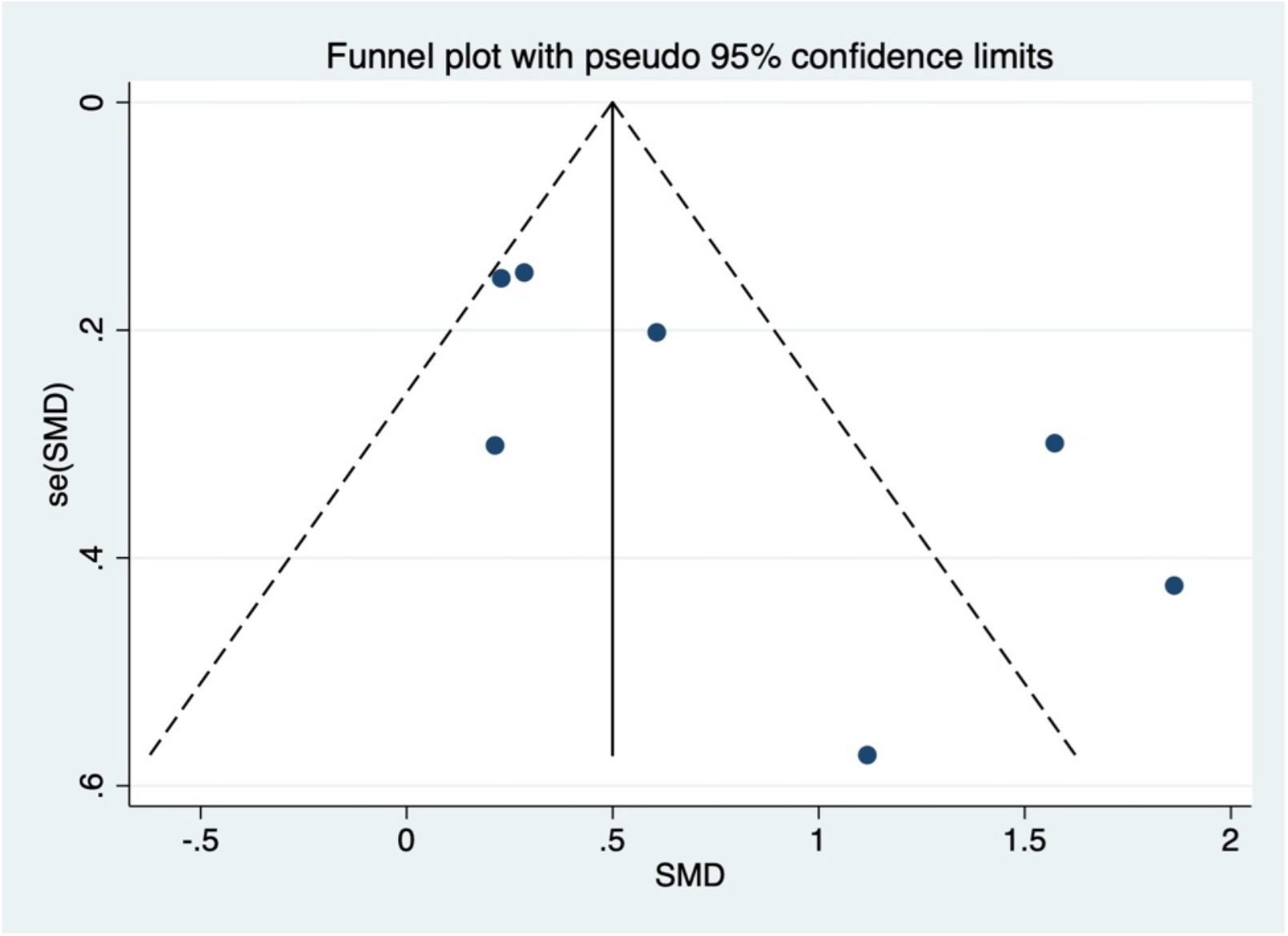
Funnel-plot analysis for the analysis of the von Willebrand Factor (VWF) antigen. SMD, standardized mean difference.

## Discussion

A single layer of healthy endothelial cells lines the entire vascular system and plays an essential role in maintaining laminar blood flow. This is attributed to its anti-inflammatory properties and non-thrombotic surface with vasodilatory homeostasis [27]. Under normal conditions, the endothelium is impermeable to large molecules, inhibits adhesion of leukocytes, provides anti-inflammatory properties, prevents thrombosis, and promotes vasodilation. Conditions that cause endothelial activation, such as infection, inflammation, or other insults, support proatherogenic mechanisms that promote plaque development by stimulating thrombin formation, coagulation, and deposition of fibrin in blood vessel walls [28].

Endothelial dysfunction is one of the manifestations of COVID-19, leading to various systemic complications through several mechanisms. Preliminary studies showed that SARS-CoV-2 directly infects endothelial cells due to the high expression of the angiotensin-converting enzyme 2 (ACE2) receptor and transmembrane protease serine 2 (TMPRSS2). After binding to SARS-CoV-2, the ACE2 receptor is internalized and downregulated, which causes a reduction of ACE2 expression in the surface of endothelial cells [2]. Inflammation in COVID-19 activates the renin-angiotensin system (RAS) either directly by increasing angiotensin I or indirectly due to the reduction of ACE2 expression on the surface of the endothelial cells. Reduced ACE2 will inhibit the hydrolysis of angiotensin II (Ang II) into angiotensin 1-7 (Ang 1-7), a vasoactive ligand of the Mas receptor (MasR), which exhibit anti-inflammatory, vasodilatory, and anti-fibrosis effects. Reduced activation of MasR will increase the Ang II type 1 receptor (AT1R) activation, thereby perpetuating the proinflammatory phenotype [29]. In addition, AT1R activation and direct viral infection of endothelial cells can increase reactive oxygen species (ROS) generation via activation of NADPH and activate nuclear factor kappa B, thereby reducing nitric oxide (NO) production [4]. Porcine animal models have recently revealed a vicious self-perpetuating pathway, whereby ROS production increases expression of ET-1, which results in further ROS production. Increased vWF expression has also been shown to increase ET-1 expression, alternatively, knockdown of vWF expression has been shown to decrease AngII-induced-ET-1 production [30]. Decreased ACE2 levels can also promote prothrombotic signaling by limiting the degradation of des-Arg9 bradykinin, thereby increasing the activated bradykinin receptors [31].

Activation of several proinflammatory cytokine receptors such as tumor necrosis factor-alpha (TNF-α) and IL-6 can directly or indirectly interfere with endothelial function. TNF-α can increase hyaluronic acid deposition in the extracellular matrix and cause fluid retention, whereas IL-6 increases vascular permeability and promote the secretion of proinflammatory cytokines by endothelial cells [32]. Subsequently, TNF-α and IL-6 induce the adhesive phenotype of endothelial cells and promote neutrophil infiltration, resulting in multiple histotoxic mediators, including neutrophil extracellular traps (NETs) and ROS to be produced, which eventually leads to endothelial cell injury. Activated endothelial cells stimulate coagulation by expressing fibrinogen, VWF, and P-selectin [32,33]. Activated endothelial by inflammatory stimuli can also induce thrombosis by favoring the expression of antifibrinolytics (e.g., PAI-1) and procoagulants (e.g., tissue factor) over the expression of profibrinolytic mediators (e.g., t-PA) and anticoagulants (e.g., heparin-like molecules and thrombomodulin) [28]. Taken together, these mechanisms contribute to massive platelet binding and formation of fibrin, leading to deposition of blood clots in the microvasculature and systemic thrombosis [2]. However, viral RNA of SARS-CoV-2 is rarely detected in the blood [34], which suggests that rather than direct viral infection of endothelial cells, additional host-dependence factors may contribute to systemic endothelial dysfunction and vasculopathy in COVID-19.

Von Willebrand factor (VWF) is a large multidomain adhesive glycoprotein produced by megakaryocytes and endothelial cells. It binds to platelet glycoproteins Ibα, αIIbβ3, and subendothelial collagen to activate platelets and initiate platelet aggregation [35]. Endothelial cell-derived VWF contributes to VWF-dependent atherosclerosis by increasing vascular inflammation and platelet adhesion and [36]. COVID-19 and endothelial activation of VWF are recognized as acute-phase proteins released from endothelial cells in response to inflammation [37], but high levels of VWF, in this case, indicated a suspicion of endothelial disturbance [38]. Expression of VWF and its release from the Weibel-Palade body of endothelial cells may also be stimulated by hypoxia. Hypoxia-induced upregulation of VWF is associated with the presence of thrombus in cardiac and pulmonary vessels that promotes leukocyte recruitment [39]. Since VWF is a significant determinant of platelet adhesion after the vascular injury leading to clot formation, high plasma levels of VWF antigen are an independent risk factor for ischemic stroke and myocardial infarction [40].

Several studies have shown that COVID-19 patients have higher plasma levels of VWF antigen than healthy controls [10,12,18,41]. The pooled means of VWF antigen levels in COVID-19 patients obtained in this study were higher than VWF levels reported in patients with acute coronary syndromes [42,43], acute ischemic stroke [44], and active ulcerative colitis [45]. Meanwhile, similar levels of VWF antigen were reported in patients with disseminated intravascular coagulation [46], severe sepsis, and septic shock not associated with COVID-19 [47,48], thus supporting the role of endothelial dysfunction in COVID-19-induced organ dysfunction. In addition to the VWF antigen, VWF activity has also been shown to increase in COVID-19 patients, thus further explaining the role of endothelial cell injury in COVID-19-associated coagulopathy [12,18,20,22].

The procoagulant state resulting from endothelial activation can also be measured from changes in the balance of tissue plasminogen activator and its endogenous inhibitor. Activated endothelial cells produce increased levels of PAI-1, which inhibits the activity of t-PA and urokinase-type plasminogen activator, causing a shift from the hemostatic balance towards the procoagulant state [49]. Increased levels of PAI-1 produced by endothelial cells and possibly other tissues, such as adipose tissue, are risk factors for atherosclerosis and thrombosis [27]. A study by Nougier *et al*. demonstrated that an impaired balance of coagulation and fibrinolysis in COVID-19 patients with significant hypercoagulability was associated with hypofibrinolysis caused by increased plasma levels of PAI-1 [26]. In COVID-19 patients, plasma levels of PAI-1 were found to be 3.7 times higher than in healthy controls [41]. A study by Kang et al. demonstrated that COVID-19 patients with severe respiratory dysfunction had significantly higher levels of PAI-1 compared to patients with burns, ARDS, and sepsis [50]. In addition to endothelial activation due to proinflammatory cytokines, direct infection by SARS-CoV-2 may cause endotheliitis, which indicates vascular endothelial damage, thereby potentiating t-PA and PAI-1 release [51].

Soluble thrombomodulin (sTM) is a soluble form in the plasma as the result of proteolytic cleavage of the intact thrombomodulin protein from the surface of endothelial cells after endothelial injury and dysfunction, such as inflammation, sepsis, ARDS, and atherosclerosis. Thrombomodulin is a vasculoprotective integral membrane type-1 glycoprotein that plays a vital role in endothelial thromboresistance [52]. The thrombomodulin-mediated binding will activate protein C, which provides anti-inflammatory, antifibrinolytic, and anticoagulant benefits for blood vessel walls [53]. In the hyperinflammatory state, increased sTM levels could be due to direct damage to the endothelial cells [54]. Elevated plasma levels of sTM have also been reported in patients with severe acute respiratory syndrome (SARS), indicating endothelial injury [55]. The plasma levels of sTM might be too low to have an impact on the coagulation process, but the consistent increase in sTM levels during pathologies has been widely considered a biomarker for endothelial dysfunction and vascular risk assessment [52]. Although the relative impact of thrombomodulin release due to endothelial injury in COVID- 19 will require further research, several studies have reported the role of sTM as a prognostic biomarker in COVID-19 patients [16,19]. These findings support the hypothesis of endotheliopathy as an important event in the transition to severity and mortality in COVID-19 patients.

### Impact for clinical practice

The elevated biomarkers of endothelial cells, which indicate a manifestation of endothelial dysfunction in COVID-19, might increase the risk of vascular complications, poor outcomes, and death. Plasma levels of VWF antigen, t-PA, PAI-1, and sTM can be used as laboratory markers for vascular risk assessment and predictors of poor outcome in COVID-19.

### Limitation

One of the 17 studies included in this meta-analysis was a preprint article. Nevertheless, a thorough assessment has been made to make sure that only sound studies are included. Most of the included studies had a retrospective observational design, and the data were not sufficiently matched or adjusted for confounders. Moreover, our primary endpoints of composite poor outcomes vary widely from ICU admission, severe illness, worsening of the respiratory status, the need for mechanical ventilation to death. Therefore, the results must be cautiously interpreted.

## Conclusion

There was an increase in the VWF antigen levels in COVID-19 patients, and the highest levels of VWF antigen were found in deceased COVID-19 patients. Biomarkers of endothelial dysfunction, including VWF antigen, t-PA, PAI-1 antigen, and sTM, are significantly associated with an increased composite poor outcome in patients with COVID-19.

## Data Availability

All data generated or analyzed during this study are included in this published article. The corresponding author (A) can be contacted for more information.

https://www.crd.york.ac.uk/prospero/display_record.php?RecordID=228821

## Data availability

The data used to support the findings of this study are included in the article and supplementary data.

## Ethics approval and consent to participate

Not Applicable.

## Consent for publication

Not Applicable.

## Acknowledgments

None.

## Supplementary data

**Table S1.**
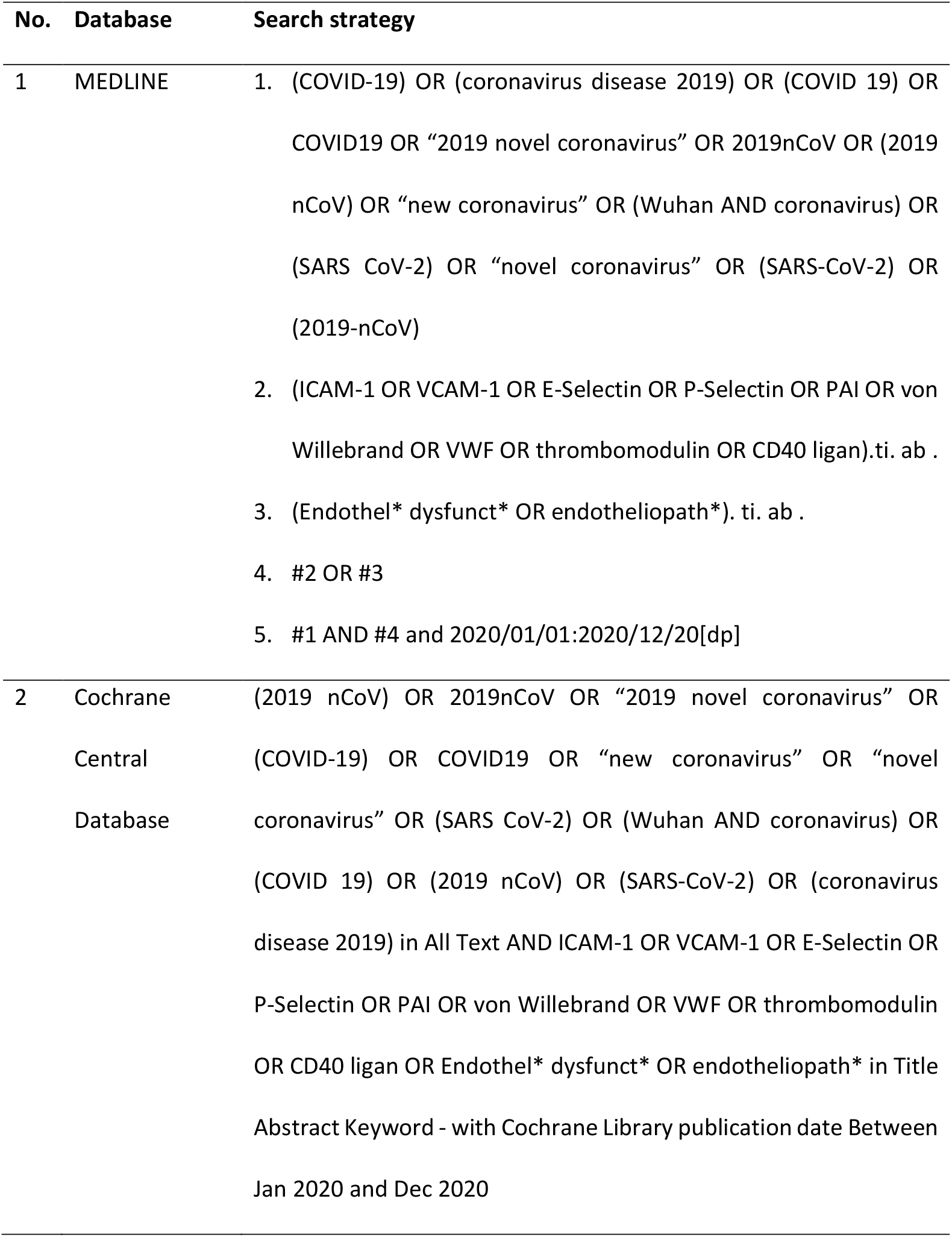
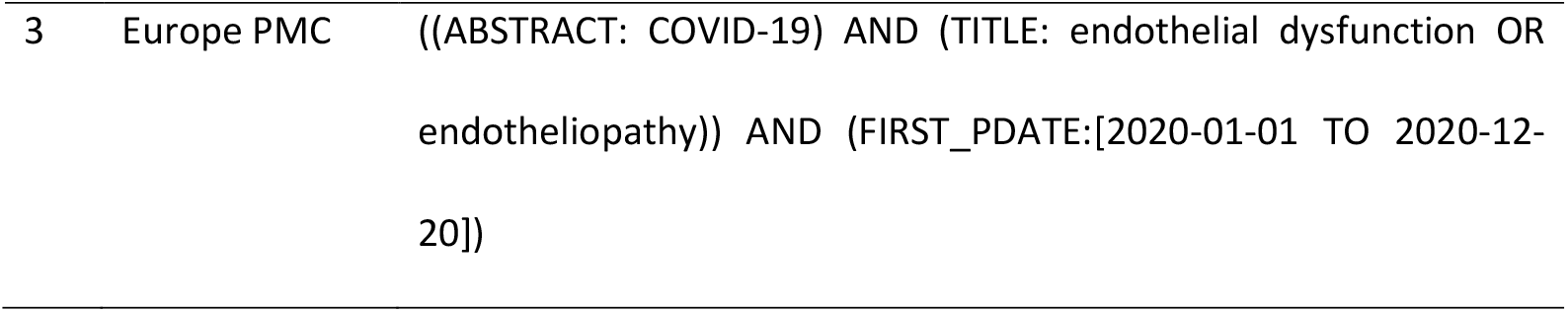
Details of the search strategy.

| No. | Database | Search strategy |
| --- | --- | --- |
| 1 | MEDLINE | 1. (COVID-19) OR (coronavirus disease 2019) OR (COVID 19) OR COVID19 OR “2019 novel coronavirus” OR 2019nCoV OR (2019 nCoV) OR “new coronavirus” OR (Wuhan AND coronavirus) OR (SARS CoV-2) OR “novel coronavirus” OR (SARS-CoV-2) OR (2019-nCoV)<br>2. (ICAM-1 OR VCAM-1 OR E-Selectin OR P-Selectin OR PAI OR von Willebrand OR VWF OR thrombomodulin OR CD40 ligan).ti. ab .<br>3. (Endothel* dysfunct* OR endotheliopath*). ti. ab .<br>4. #2 OR #3<br>5. #1 AND #4 and 2020/01/01:2020/12/20[dp] |
| 2 | Cochrane Central Database | (2019 nCoV) OR 2019nCoV OR “2019 novel coronavirus” OR (COVID-19) OR COVID19 OR “new coronavirus” OR “novel coronavirus” OR (SARS CoV-2) OR (Wuhan AND coronavirus) OR (COVID 19) OR (2019 nCoV) OR (SARS-CoV-2) OR (coronavirus disease 2019) in All Text AND ICAM-1 OR VCAM-1 OR E-Selectin OR P-Selectin OR PAI OR von Willebrand OR VWF OR thrombomodulin OR CD40 ligan OR Endothel* dysfunct* OR endotheliopath* in Title Abstract Keyword - with Cochrane Library publication date Between Jan 2020 and Dec 2020 |
| 3 | Europe PMC | ((ABSTRACT: COVID-19) AND (TITLE: endothelial dysfunction OR<br>endotheliopathy)) AND (FIRST_PDATE:[2020-01-01 TO 2020-12-20]) |

**Table S2.** Assessment of included studies in the Newcastle-Ottawa Scale (NOS).

| No | Study | Selection<br>(max 4 stars) | Comparability<br>(max 2 stars) | Outcome<br>(max 3 stars) |
| --- | --- | --- | --- | --- |
| 1 | Cugno, 2020 | **** | * | *** |
| 2 | Fan, 2020 | *** | * | ** |
| 3 | Goshua, 2020 | **** | ** | *** |
| 4 | Helms, 2020 | *** | ** | ** |
| 5 | Henry, 2020 | **** | ** | *** |
| 6 | Hoechter, 2020 | *** | ** | ** |
| 7 | Ladikou, 2020 | *** | ** | ** |
| 8 | Mancini, 2020 | *** | * | *** |
| 9 | Morici, 2020 | *** | * | ** |
| 10 | Nougier, 2020 | **** | ** | *** |
| 11 | Panigada, 2020 | *** | * | ** |
| 12 | Pine, 2020 | *** | ** | *** |
| 13 | Ranucci, 2020 | *** | ** | *** |
| 14 | Rauch, 2020 | **** | * | ** |
| 15 | Rovas, 2020 | **** | ** | *** |
| 16 | Sweeney, 2020 | **** | * | ** |
| 17 | Taus, 2020 | **** | ** | ** |

